# Prevalence of differences of sex development in children and adolescents in Switzerland from 2000-2019

**DOI:** 10.1101/2024.03.11.24304115

**Authors:** Sara Metzger, Grit Sommer, Christa E. Flück, Swiss DSD Cohort Study Group

**Affiliations:** Division of Pediatric Endocrinology, Diabetology and Metabolism, Department of Pediatrics, Inselspital, Bern University Hospital, University of Bern, Switzerland; Department of Biomedical Research, University of Bern, Switzerland; Institute of Social and Preventive Medicine, University of Bern, Switzerland

**Author notes:** co-last authors (Grit Sommer, Christa E. Flück). Members of the Swiss DSD Cohort Study Group in alphabetical order: Christine Aebi-Ochsner, Private Practice for Paediatric Endocrinology, Biel Switzerland; Kanetee Busiah, Department of Pediatric Endocrinology and Diabetology, CHUV, University Children’s Hospital, Lausanne, Switzerland; Mirjam Dirlewanger, Pediatric Endocrine and Diabetes Unit, Children’s University Hospital Geneva, Geneva, Switzerland; Sylvia Gschwend, Private Practice for Paediatric Endocrinology, Zug, Switzerland; Melanie Hess, Pediatric Endocrinology and Diabetology, University Children’s Hospital Basel, Basel, Switzerland; Beatrice Kuhlmann, Pediatric Endocrinology, Cantonal Hospital Aarau, Aarau, Switzerland and Private Practice for Paediatric Endocrinology, Basel, Switzerland; Dagmar l’Allemand, Department of Endocrinology, Children’s Hospital of Eastern Switzerland, St. Gallen, Switzerland; Mariarosaria Lang, Paediatric Endocrinology and Diabetology, University Children’s Hospital Zurich, Zurich, Switzerland; Kees Noordam, Centre for Paediatric Endocrinology Zurich (PEZZ), Zurich, Switzerland; Franziska Phan-Hug, Department of Pediatrics, Hospital Morges, Morges and Department of Pediatrics, Hospital Wallis, Sion, Switzerland; Ursina Probst, Department of Pediatrics, Cantonal Hospital Winterthur, Winterthur, Switzerland; Maristella Santi, Department of Pediatrics, Cantonal Hospital Lucerne, Lucerne, Switzerland; Silvia Schmid, Private Practice for Paediatric Endocrinology, Dubendorf, Switzerland; Valérie Schwitzgebel, Pediatric Endocrine and Diabetes Unit, Children’s University Hospital Geneva, Geneva, Switzerland; Michael Steigert, Department of Pediatrics, Cantonal Hospital Graubuenden, Chur, Switzerland; Gabor Szinnai, Pediatric Endocrinology and Diabetology, University Children’s Hospital Basel, Basel, Switzerland; Gerald Theintz, Private Practice for Paediatric Endocrinology, Lausanne, Switzerland. **Correspondence:** Grit Sommer Institute of Social and Preventive Medicine University of Bern Mittelstrasse 43 3012 Bern, Switzerland.

**Keywords:** DSD, intersex, sex chromosome DSD, 46, XY DSD, 46, XX DSD, CAH, gonadal development

## Abstract

**Objective:** Reliable data on prevalence of differences of sex development (DSD) are lacking. We aimed to estimate population-based prevalence of DSD in Switzerland.

**Design:** Retrospective population-based study including children and adolescents with DSD according to Chicago Consensus, born in Switzerland from 2000-2019.

**Methods:** Endocrine care centers in ten Swiss Children’s Hospitals and eight private endocrine practices collected DSD data through the I-DSD registry or case report forms. We calculated prevalence for DSD diagnostic groups and analyzed trends in prevalence.

**Results:** Over the 20-year study period, we identified 561 individuals with DSD. Almost half (n=266, 47%) had sex chromosome DSD, 177 (32%) had 46,XY DSD and 118 (21%) had 46, XX DSD. Causes for 46,XY DSD were disturbed androgen synthesis or action (37/177, 21%), atypical gonadal development (28/177, 16%), or other causes (112/177, 63%). Causes for 46,XX DSD were androgen excess (99/118, 84%), atypical gonadal development (8/118, 7%), or other causes (11/118, 9%). On average, 28 new cases were born with DSD annually. Prevalence was 17 for sex chromosome DSD, 12 for 46,XY DSD and 8 for 46,XX DSD per 100’000 live births and year. One per 7’500 newborn girls had 46,XX congenital adrenal hyperplasia (CAH).

**Conclusion:** Prevalence of sex chromosome DSD was lower than expected because of underreporting due to late diagnosis. Prevalence of 46,XX CAH is similar to newborn screening data, suggesting good completeness of cases. For complex DSD cases, we expect complete coverage. This study provides a valuable resource for policymaking and (inter)national research on DSD.

## INTRODUCTION

Differences of sex development (DSD) is an umbrella term that stands for a heterogeneous group of rare conditions that affect human sex development and maturation (1). The term DSD has first been introduced with the Chicago Consensus Statement in 2006 that defined DSD as "congenital conditions in which the development of chromosomal, gonadal, anatomic sex is atypical" (2). Most DSDs are congenital due to their genetic origins. However, many individuals never receive a genetic diagnosis but are diagnosed as DSD based on their phenotype (3).

Although this consensus around terminology has been widely accepted by medical professionals, it remains controversial (4), and other terms have been suggested including intersex, variations of sex development or more recently variations of sex characteristics. New knowledge on human sex development and the introduction of a classification based on genetics clarified the perspective on this broad and complex subject. A clear definition allowed researchers and physicians to identify and work together with persons with DSD and improve efforts towards well-integrated, progressive, patient-centered care and research across the DSD spectrum. The International Registries For Rare Conditions Affecting Sex Development & Maturation (SDMregistries) include international databases for DSD (e.g., the I-DSD Registry) that allow for standardized and pseudonymized data collection (5). They provide valuable tools for researchers and physicians to record and obtain data of individuals with DSD to solve clinical and translational research questions (5, 6). As of June 2024, the I-DSD Registry included data of 9165 individuals with DSD originating from 152 centers of 46 countries world-wide (7). It has formed the basis of over 65 research projects addressing various topics of DSD such as mechanisms of disease of specific subtypes of DSD, genetic and diagnostic approaches, treatment options, long-term outcomes, quality of care and psychosocial topics.

Recent developments have shifted perceptions and approaches toward DSD, yet many open research questions remain to improve care for individuals with DSD (1, 8–10). Provision of optimal healthcare for individuals with DSD and perception by policymakers, ethical and legal bodies as well as society require a clear understanding of the occurrence of these conditions. However, the accurate incidence and prevalence of DSD are still unknown (11). One exception are studies reporting incidences of congenital adrenal hyperplasia (CAH), because CAH is included in national neonatal screening programs (12). Only few studies estimated incidence or prevalence for other types of DSD, but these studies were either old, dating from 1955 to 2000 (13), or covered only single centers or regions and thus yielded variable results (14) (15–23). To date, no country has a nation-wide registration system for DSD.

We aimed to calculate prevalence and trends of DSD diagnostic groups according to the Chicago Consensus Classification in children and adolescents born population based from 2000-2019 in Switzerland.

## MATERIAL AND METHODS

### Ethics, Study Design and Participants

For this population-based retrospective observational study cohort, we retrospectively identified patients with DSD in 18 different Swiss centers of pediatric endocrinology.

We included all individuals with DSD as defined by the Chicago Consensus Classification (2), who were born in Switzerland between January 1, 2000 and December 31, 2019. In Switzerland, care for children and adolescents with DSD is mainly provided by a specialized interdisciplinary DSD team (4) at outpatient clinics of pediatric hospitals under the lead of a pediatric endocrinologist. Additionally, there are pediatric endocrinologists in private practices that may provide care for a few individuals, predominantly with Turner syndrome, Klinefelter syndrome or CAH. We approached all 11 departments of pediatric endocrinology in pediatric hospitals to collaborate for data collection, and 10 out of 11 participated covering the whole of Switzerland except the canton of Ticino. We also contacted the pediatric endocrinologists in private practices to participate in our study via email through the Swiss Society of Pediatric Endocrinology and reminded them if we did not hear back. Of a total of 16 private practices, we heard back from 13, of which 5 did not treat eligible children and adolescents. The remaining 3 who did not answer are small practices.The Ethics Committee of the Canton of Bern granted ethical approval to the Swiss DSD Cohort Study under BASEC ID 2016-01210. With ethical approval, we were able to collect a core data set without informed consent of participants.

### Data Collection

Participating centers identified eligible patients through patient lists of treating physicians and searches in their administrative clinical databases or hospital coding systems. Physicians extracted data from medical records and filled them into the basic registration module of the I-DSD Registry (https://sdmregistries.org/). Centers without access to the registry used a case report form identical to the variables in the I-DSD Registry. Extracted data included treatment center, year of birth, assigned sex at birth, country of birth, karyotype, details of DSD condition including genetic analyses and current gender. We coded each individual according to the diagnostic categories suggested from the Chicago Consensus Classification (2) into three DSD main groups (sex chromosome DSD, 46,XY DSD, 46,XX DSD) and diagnostic subgroups (sex chromosome DSD: Klinefelter syndrome and variant, Turner syndrome and variants, 45,X/46,XY and variants, other; 46,XY DSD: disorders of gonadal development, disorders in androgen synthesis or action, other (including severe, complex hypospadias); 46,XX DSD: disorders of gonadal development, androgen excess (including 46,XX CAH), other. We clarified missing or inconclusive data with the collaborating physicians.

Population-based data on live births came from the Swiss Federal Office of Statistics (SFSO). Numbers of cases of CAH identified by newborn screening came from the Swiss Neonatal Screening laboratory (https://www.neoscreening.ch/de/).

To ensure that our cohort did not include duplicate entries of the same individuals with DSD, an independent institution performed record linkage based on first and last name and date of birth.

### Data Analysis and Statistics

We calculated prevalence based on the day the individuals were born, assuming that any DSD regardless of the timing of clinical presentation, is already present at birth, similarly to a Danish study on CAH (24). We calculated prevalence based on population data on live births from the Swiss Federal Statistical Office (SFSO). The SFSO publishes live birth statistics by year and canton, which allowed us to exclude population data of Ticino (25). We calculated the average number of cases born with DSD per year for main DSD categories and specific DSD subgroups during the entire observation period from 2000-2019 and for four 5-year intervals (2000–2004, 2005–2009, 2010–2014, 2015–2019). We calculated prevalence of DSD expressed per 100’000 liveborn individuals per year. Depending on the diagnosis, we calculated prevalence based on all newborns, or only newborn boys or newborn girls. For all DSDs combined, for the main DSD categories (sex chromosome DSD, 46,XY DSD, 46,XX DSD), and for two sex chromosome DSD subgroups (45,X/46,XY and variants, other sex chromosome DSD) we used the number of all newborns. For Klinefelter syndrome and variants and the 46,XY DSD subgroups (disorders of gonadal development, disorders in androgen synthesis or action, other 46,XY DSD) we used newborn boys. For Turner syndrome and variants and the 46,XX DSD subgroups (disorders of gonadal development, androgen excess, other 46,XX DSD) we used newborn girls. We calculated confidence intervals for prevalence using the Poisson distribution (26). We used Stata, Version 16.1 for data preparation and descriptive statistics, and R statistic management, R Version 4.2.2 for prevalence calculations.

To examine trends in prevalence expressed as annual percentage changes (APC), we used JoinPoint, Version 4.0.2.2. JoinPoint is a statistical package originally developed by the US National Cancer Institute to describe trends in cancer incidence and mortality (27). JoinPoint fitted regression lines through the selected data, with the natural logarithm of proportion (prevalence) as dependent variable and calendar year (2000–2019) as independent variable. We allowed for a maximum number of 0 joinpoints to capture overarching trends within the data.

## RESULTS

A total of 685 individuals with DSD born between 2000 and 2019 were treated in the 10 pediatric hospitals including all University Children’s Hospitals in Switzerland and the 8 private pediatric practices. We excluded duplicate individuals (n=56) and individuals born outside of Switzerland or in the canton Ticino (n=66), leading to a final cohort of 561 individuals with DSD (**Figure 1**).

**Figure 1:**
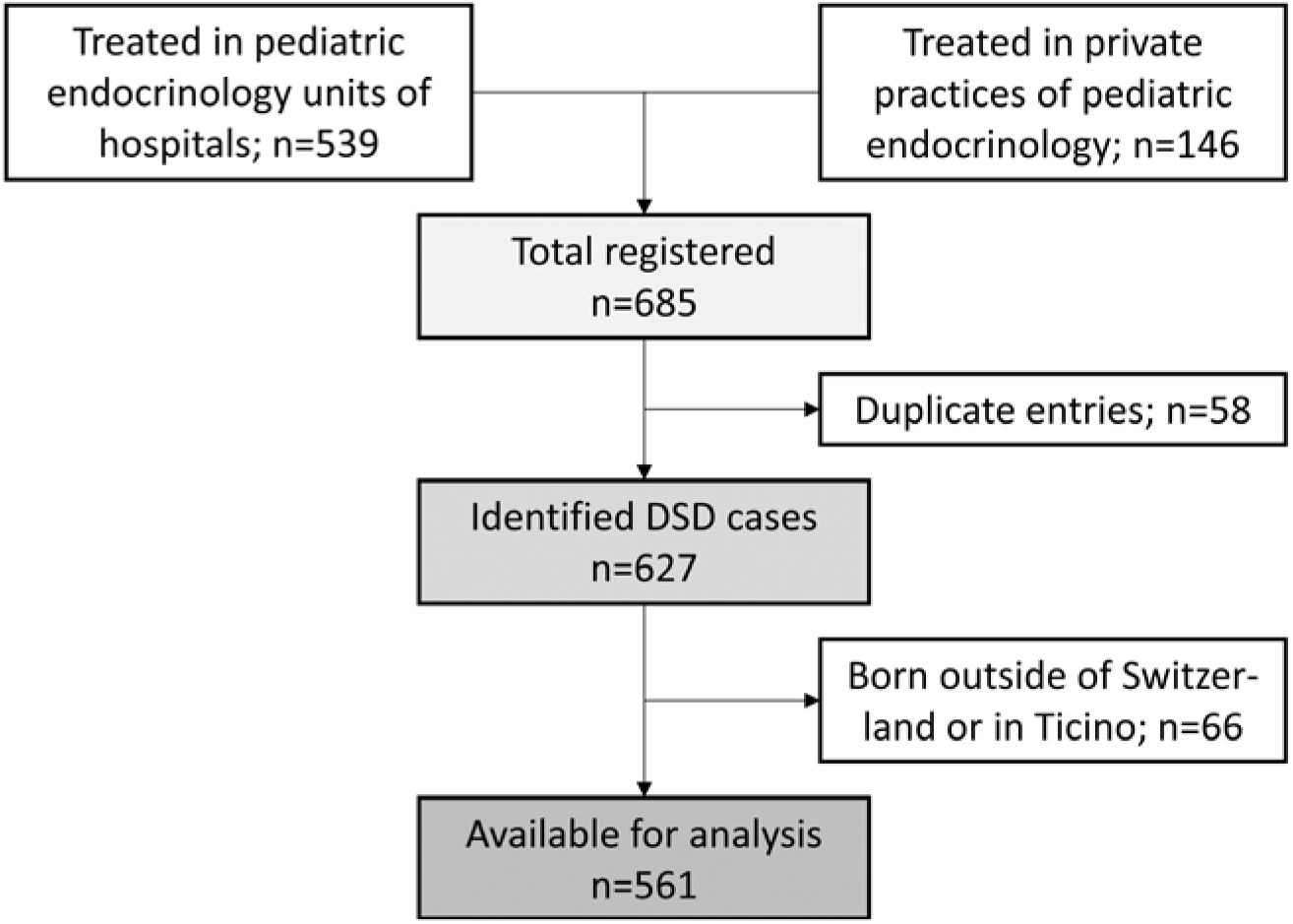
Identification of the Swiss DSD cohort. Study participants were identified through pediatric endocrinology departments in pediatric hospitals and private practices specialized in endocrine care.

We classified our cohort of 561 individuals into the three main DSD categories according to the Chicago Consensus Classification (2). A total of 266 (47%) individuals had sex chromosome DSD, 177 (32%) 46,XY DSD and 118 (21%) 46,XX DSD. **Table 1** gives an overview of the specific diagnoses comprised within these three main DSD categories.

**Table 1:** Numbers and proportions of DSD cases in Switzerland born between 2000 and 2019, by sex at birth.

|  | Total |  | Sex at birth |  |  |  |  |  |
| --- | --- | --- | --- | --- | --- | --- | --- | --- |
|  |  |  | Female |  | Male |  | Other/<br>Not assigned |  |
|  | N=561 |  | N=301 |  | N=242 |  | N=18 |  |
|  | n | % | n | % | n | % | n | % |
| **Period of birth** |  |  |  |  |  |  |  |  |
| 2000-2004 | 146 | 26 | 83 | 28 | 61 | 25 | 2 | 11 |
| 2005-2009 | 160 | 28 | 92 | 31 | 60 | 25 | 8 | 44 |
| 2010-2014 | 136 | 24 | 78 | 26 | 54 | 22 | 4 | 22 |
| 2015-2019 | 119 | 21 | 48 | 16 | 67 | 28 | 4 | 22 |
| **Sex chromosome DSD** | 266 | 100 | 171 | 100 | 95 | 100 | 0 | n/a |
| Klinefelter syndrome and variants | 74 | 28 | 0 | 0 | 74 | 78 | 0 | n/a |
| Turner syndrome and variants | 149 | 56 | 148 | 87 | 1 | 1 | 0 | n/a |
| 45,X/46,XY and variants | 33 | 12 | 22 | 13 | 11 | 12 | 0 | n/a |
| Other | 10 | 4 | 1 | 1 | 9 | 9 | 0 | n/a |
| **46,XY DSD** | 177 | 100 | 28 | 100 | 135 | 100 | 14 | 100 |
| *Disorders of gonadal development* | 28 | 16 | 9 | 32 | 17 | 13 | 2 | 14 |
| Complete or partial gonadal dysgenesis | 16 |  | 8 |  | 7 |  | 1 |  |
| Gonadal regression | 9 |  | 0 |  | 9 |  | 0 |  |
| Ovotesticular DSD | 3 |  | 1 |  | 1 |  | 1 |  |
| *Disorders in androgen synthesis or action* | 37 | 21 | 18 | 64 | 18 | 13 | 1 | 7 |
| Disorders of androgen synthesis | 12 |  | 2 |  | 9 |  | 1 |  |
| Disorders of androgen action | 19 |  | 16 |  | 3 |  | 0 |  |
| Disorders of AMH or AMH receptor | 6 |  | 0 |  | 6 |  | 0 |  |
| *Other* | 112 | 63 | 1 | 4 | 100 | 74 | 11 | 79 |
| Complex genital anomalies<sup>a</sup> | 106 |  | 1 |  | 95 |  | 10 |  |
| Cloacal anomalies | 2 |  | 0 |  | 1 |  | 1 |  |
| Other | 4 |  | 0 |  | 4 |  | 0 |  |
| **46,XX DSD** | 118 | 100 | 102 | 100 | 12 | 100 | 4 | 100 |
| *Disorders of gonadal development* | 8 | 7 | 2 | 2 | 6 | 50 | 0 | 0 |
| Gonadal dysgenesis | 2 |  | 2 |  | 0 |  | 0 |  |
| Ovotesticular DSD | 2 |  | 0 |  | 2 |  | 0 |  |
| Testicular DSD | 4 |  | 0 |  | 4 |  | 0 |  |
| *Androgen excess* | 99 | 84 | 89 | 87 | 6 | 50 | 4 | 100 |
| Fetal<sup>b</sup> | 97 |  | 87 |  | 6 |  | 4 |  |
| Fetoplacental | 2 |  | 2 |  | 0 |  | 0 |  |
| Maternal | 0 |  | 0 |  | 0 |  | 0 |  |
| *Other* | 11 | 9 | 11 | 11 | 0 | 0 | 0 | 0 |
| Cloacal anomalies | 7 |  | 7 |  | 0 |  | 0 |  |
| Other | 4 |  | 4 |  | 0 |  | 0 |  |
| **Gender at last follow-up** |  |  |  |  |  |  |  |  |
| Female | 304 | 54 | 294 | 52 | 5 | 1 | 5 | 1 |
| Male | 256 | 46 | 6 | 1 | 237 | 42 | 13 | 2 |
| Both | 1 | 0.2 | 1 | 0.2 | 0 | 0 | 0 | 0 |

Abbreviations: AMH, Anti-Mullerian hormone; DSD, differences of sex development; n/a, not applicable.
<sup>a</sup>Complex genital anomalies in 46,XY DSD include severe hypospadias (penoscrotal).
<sup>b</sup>Fetal androgen excess in 46,XX DSD includes congenital adrenal hypoplasia (CAH).

### Prevalence of DSD in Switzerland

Throughout the entire 20-year study period spanning from 2000 to 2019, an annual average of 76’719 children were born in all Swiss cantons excluding Ticino. During the same period, we identified an average number of 28 individuals with DSD born per year (**Table 2**), resulting in an overall prevalence of 36.6 (95%CI 33.6; 39.7) per 100’000 newborns per year for all DSD diagnoses (**Table 2**).

**Table 2.** Prevalence, average number of cases per year of individuals identified with DSD in Switzerland, born between 2000-2019, by diagnostic group.

|  | **Average**<br>**number of**<br>**cases/year** | **Prevalence per**<br>**100.000**<br>**newborns per**<br>**year (95% CI)** | **1:N**<br>**newborns** |
| --- | --- | --- | --- |
| *All DSD diagnoses<sup>a</sup>* | 28.1 | 36.6 (33.6; 39.7) | 2735 |
| *DSD diagnostic group* |  |  |  |
| Sex chromosome DSD<sup>a</sup> | 13.3 | 17.3 (15.3; 19.5) | 5768 |
| 46,XY DSD<sup>a</sup> | 8.9 | 11.5 (9.9; 13.4) | 8669 |
| 46,XX DSD<sup>a</sup> | 5.9 | 7.7 (6.4; 9.2) | 13003 |
| *Sex chromosome DSD* |  |  |  |
| Klinefelter syndrome and variants<sup>b</sup> | 3.7 | 9.4 (7.4; 11.8) | 10660 |
| Turner syndrome and variants<sup>c</sup> | 7.5 | 20.0 (16.9; 23.5) | 5004 |
| 45,X/46,XY and variants<sup>a</sup> | 1.7 | 2.2 (1.5; 3.0) | 46496 |
| Other<sup>a</sup> | 0.5 | 0.7 (0.3; 1.2) | 153437 |
| *46,XY DSD* |  |  |  |
| Disorders of gonadal development<sup>b</sup> | 1.4 | 3.5 (2.4; 5.1) | 28172 |
| Disorders in androgen synthesis or action<sup>b</sup> | 1.9 | 4.7 (3.3; 6.5) | 21319 |
| Other<sup>b</sup> | 5.6 | 14.2 (11.7; 17.1) | 7043 |
| *46,XX DSD* |  |  |  |
| Disorders of gonadal development<sup>c</sup> | 0.4 | 1.1 (0.5; 2.1) | 93195 |
| Androgen excess<sup>c</sup> | 5.0 | 13.3 (10.8; 16.2) | 7531 |
| Other<sup>c</sup> | 0.6 | 1.5 (0.7; 2.6) | 67778 |

Abbreviations: CI, confidence interval; DSD, differences of sex development.
<sup>a</sup>Prevalence calculated based on newborns of both sexes.
<sup>b</sup>Prevalence calculated based on newborn boys.
<sup>c</sup>Prevalence calculated based on newborn girls.

When we stratified our study period into 5-year intervals, annual DSD prevalence per 100’000 newborns was highest with 44.3 (95%CI 31.7; 51.7) between 2005-2009, and lowest with 28.2 (95%CI 23.4; 33.7) in the most recent interval between 2015-2019 (Figure 2A and Supplemental Table 1 https://boris-portal.unibe.ch/handle/20.500.12422/33491). Sex chromosome DSD accounted for the largest proportion of cases among all DSD main groups between 2000-2014. Only between 2015-2019, the 46,XY DSD proportion was larger. Throughout all time intervals, the 46,XX DSD group accounted for the smallest proportion of cases.

**Figure 2:**
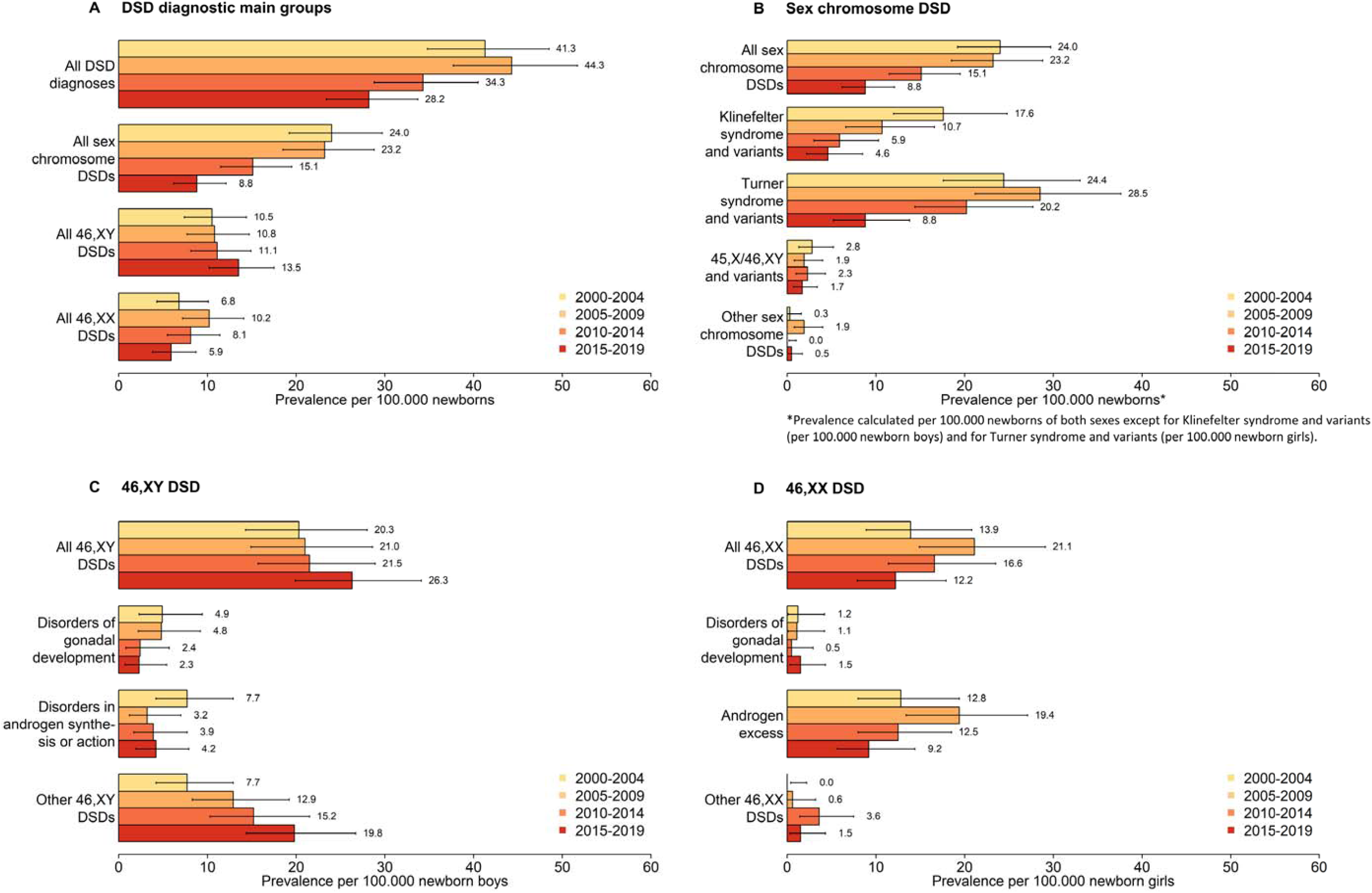
Comparison of prevalence of identified individuals with DSD in Switzerland between 2000-2004, 2005-2009, 2010-2014 and 2015-2019, by DSD diagnostic group, according to the Chicago Consensus Classification. Error bars indicate 95% confidence intervals. Abbreviations: DSD, differences of sex development.

Sex chromosome DSD prevalence was highest with 24.0 (95%CI 19.2; 29.7) per 100’000 newborns per year between 2000-2004 (Figure 2B and Supplemental Table 1 https://boris-portal.unibe.ch/handle/20.500.12422/33491). Turner syndrome and variants accounted for the largest proportion of sex chromosome DSD (prevalence 20.0 per 100’000 newborn girls, 95%CI 16.9;23.5), followed by Klinefelter syndrome and variants (prevalence 9.4 per 100’000 newborn boys, 95%CI 7.4-11.8). Other types of sex chromosome DSD were rare (prevalence 0.5-1.7 per 100’000 newborns). Prevalence of sex chromosome DSD decreased from 2000-2019 with an APC of -6.3 (95%CI -8.4; -4.1) which was mainly driven by the Klinefelter syndrome (APC -8.2, 95%CI 11.4; -4.8) and Turner syndrome group (APC -5.2, 95%CI -7.8; -2.5) (Figures 2B, Supplemental Figure 1B https://boris-portal.unibe.ch/handle/20.500.12422/33491).

Prevalence of 46,XY DSD was 11.5 (95%CI 9.9; 13.4) per 100’000 newborns for the entire study period and remained stable over time (APC 1.8, 95%CI -0.5; 4.1) (Table 2, Supplemental Figure 1C https://boris-portal.unibe.ch/handle/20.500.12422/33491), with a slight trend towards an increasing prevalence after 2014 (Figure 2C). Other 46,XY DSD (prevalence 14.2 per 100’000 newborn boys, 95%CI 11.7; 17.1) was highest among the group of 46,XY DSD, followed by disorders in androgen synthesis or action (prevalence 4.7 per 100’000 newborn boys, 95%CI 3.3; 6.5) and disorders of gonadal development (prevalence 3.5 per 100’000 newborn boys, 95%CI 2.4; 5.1) (Table 2). While prevalence of other 46,XY DSD increased over time (APC 4.5, 95%CI 1.0-8.1), prevalence of disorders of gonadal development or disorders in androgen synthesis or action remained stable (Supplemental Figure 1C https://boris-portal.unibe.ch/handle/20.500.12422/33491).

Prevalence of 46,XX DSD was 7.7 (95%CI 6.4; 9.2) per 100’000 newborns (Table 2, Figure 2D), and remained stable over time (APC -2.3, 95%CI -5.9; 1.4) (Table 2, Supplemental Figure 1D https://boris-portal.unibe.ch/handle/20.500.12422/33491). Among 46,XX DSD, the androgen excess group had the highest prevalence with 13.3 (95%CI 10.8; 16.2) per 100’000 newborn girls with stable prevalence over the study period. Other types of 46, XX DSD were rare (prevalence 1.1-1.5 per 100’000 newborn girls).

### Sex registration at birth

In Switzerland sex registration at birth is mandatory and only the traditional categories of either male or female are officially available. We did not observe a difference in ratio of sex registration over the study period (Table 1). In our cohort of 561 individuals, 301 (54%) were registered with female sex at birth and 242 (43%) with male sex at birth. For 18 individuals, physicians reported that sex directly at birth was not assigned (3%; n=14 46,XY and n=4 46,XX), and data on official sex registration at birth was not available to us. All individuals with sex chromosome DSD were registered with a female or male sex at birth. Discordance between karyotype and sex registration at birth was found in 28/177 individuals with 46,XY DSD (16%) and in 12/118 individuals with 46,XX DSD (10%). A difference between registered sex at birth and gender at last follow-up was reported in 5 individuals originally registered female at birth (1%) and in 7 individuals originally registered male at birth (1%) with one individual identifying as both male and female.

### Where are individuals with DSD cared for?

We found that most children and adolescents with DSD in Switzerland (452/561, 80%) were seen by pediatric endocrinologists working in hospitals with a interdisciplinary DSD team (Figure 3). Pediatric endocrinologists in private practice cared predominantly for children and adolescents with Turner syndrome, Klinefelter syndrome, or 46,XX androgen excess.

**Figure 3:**
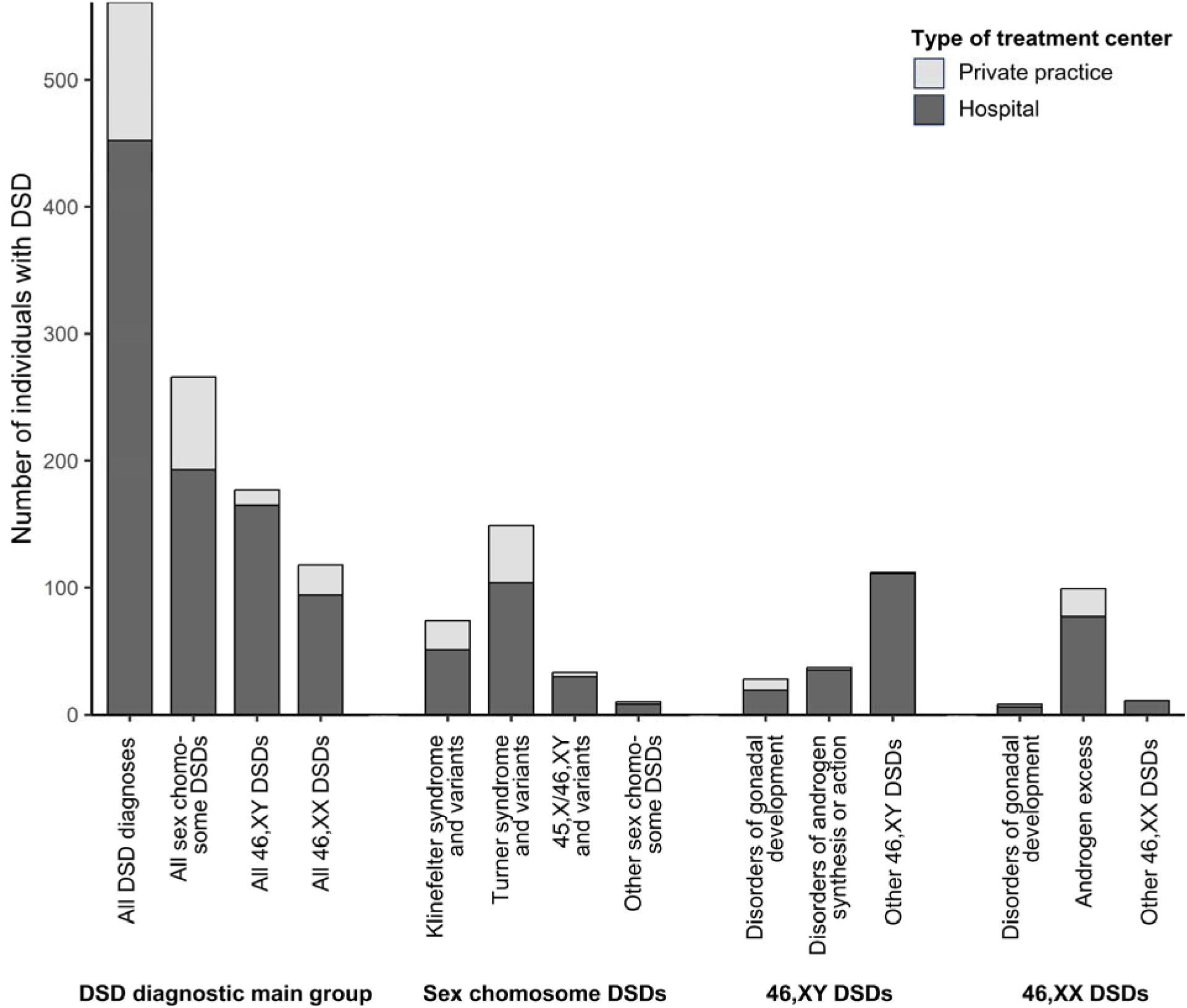
Number of individuals with DSD according to the Chicago Consensus Classification by type of treatment center, identified in Switzerland between 2000-2019. Abbreviations: DSD, differences of sex development.

## DISCUSSION

We report prevalence for different DSD diagnoses according to the Chicago Consensus Classification in a population-based cohort between 2000-2019 in Switzerland. Prevalence of DSD treated by pediatric endocrinologist was 37 per 100’000 (1 of 2’735) newborns including sex chromosome, 46,XY and 46,XX DSD. Overall, DSD prevalence decreased from 2000-2019, which was driven by a strong decrease in prevalence of sex chromosome DSD. Prevalence of 46,XY DSD tended to increase and prevalence of 46,XX DSD to decrease over time.

Most studies on epidemiology of DSD come from single centers in different countries. These studies mostly describe prevalence of DSD (15, 18, 20, 21, 28) or atypical genitalia (16, 19, 22), and only some used the Chicago Consensus Classification. The reported prevalence at birth ranges from 280 per 100’000 (1 of 357) when including any kind of atypical genitalia (28) to 16 per 100’000 (1 of 6’347) when using DSD diagnoses according to the Chicago Consensus Classification (21). The two existing multicenter studies from the US (17) and Scotland (23) reported numbers of children with either atypical genitalia or DSD at birth. The Scottish study registered atypical genitalia in Scotland prospectively between 2013-2019. They found that 53 per 100’000 (1 of 1’881) newborns, the treating physicians suspected a DSD and 30 per 100’000 (1 of 3’318) newborns required DSD specialist consultation (23). The US study by Finlayson et al (17) compared relative prevalence of DSD diagnoses within a cohort of 99 infants with atypical genitalia to other cohorts and found large differences in relative prevalence of sex chromosome, 46,XY and 46,XX DSD between cohorts. Several factors may contribute to this broad variability, including definition of diagnosis (e.g. inclusion/exclusion of cases without atypical genitalia), differences in study population (e.g. ethnicity, consanguinity), sampling (single center versus multicenter), study design (e.g. prospective, cross-sectional), age of study participants (e.g. newborns, children, all ages). Thus population-based data to obtain representative estimates of prevalence for different DSD diagnoses is crucial.

### Sex chromosome DSD

We calculated prevalence of DSD based on live birth rates in Switzerland. Our overall prevalences are lower than published data for sex chromosome DSD (29, 30). Turner syndrome occurs in 25-50 individuals per 100’000 females (29, 31). We found a prevalence of 24-29 per 100’000 newborn girls for Turner syndrome and variants during 2000-2009, which decreased to 9 per 100’000 during 2015-2019. The reason for this decrease is because less than one third of individuals with Turner syndrome are diagnosed in childhood or adolescence (29, 31). The reported prevalence for Klinefelter syndrome varies from 40-250 per 100’000 males with the main proportion only diagnosed in adulthood (32, 33). For Klinefelter and variants we found a prevalence of 18 per 100’000 newborn boys during 2000-2004 that decreased to 5 per 100’000 during 2015-2009. Our cohort included individuals with DSD who were born from 2000-2019, thus a large proportion of individuals were too young to have received a Turner syndrome or Klinefelter syndrome diagnosis, for example because they were still prepubertal at time of study, or because fertility concerns may arise only later in adulthood. Cohort studies with systematic screening or longer observation periods are needed to capture real prevalence estimates for individuals with Turner and Klinefelter syndrome. Some individuals with Turner or Klinefelter syndrome may not have required pediatric endocrine care and thus were missed by our cohort study. Pregnancy termination after a prenatal diagnosis of sex chromosome DSD leads to additional underestimation of sex chromosome prevalence (34). We only included data of live births, thus we cannot determine to what extent this would affect our prevalence estimates.

### Non-chromosomal DSD

We assume almost complete coverage of rare and complex subgroups of non-chromosomal DSD with atypical genitalia. These individuals are mostly diagnosed at birth and require extensive diagnostic assessments which leads to referral to interdisciplinary DSD teams at pediatric hospitals. Specialized DSD physicians of these interdisciplinary teams reported all their DSD cases to our study cohort, reducing the likelihood of missing eligible individuals. Comparable prevalence data is scarce for individuals with complex forms of DSD.

46,XY DSD comprise disorders of gonadal development, disorders in androgen synthesis or action, and other forms. A Swedish study (35) found a prevalence of 6.4 per 100’000 live born females for individuals with a 46,XY karyotype and a female phenotype. These figures are hard to compare to our results, because we calculated prevalence based on newborn boys and did not include data on phenotype. We observed a prevalence of 3.5 per 100’000 newborn boys for individuals with disorders of gonadal development and of 4.7 for individuals with androgen synthesis or action. Our estimates for prevalence of 46,XY individuals with disorders of androgen sensitivity or action likely underestimated the true prevalence, because those with a female phenotype may perhaps only be evaluated after pubertal age due to primary amenorrhea. The largest group among the 46,XY DSD was the heterogeneous group of other 46,XY DSD including complex genital and cloacal anomalies. Often individuals of this group are not classified as DSD in hospital administration coding systems, as they have their first presentation in departments other than endocrinology (e.g. urology). Only a fraction is referred to specialized DSD teams for interdisciplinary care and genetic work-up. Genetic work-up does not yield a genetic diagnosis relating to a DSD in >50% of individuals (36). In a US single center study among 131 boys with proximal hypospadias, only 60 (46%) received endocrine and genetic testing and 9 (7%) received a DSD diagnosis (37). In a Chinese single center study in 165 individuals with proximal hypospadias, only 14 (8%) obtained a DSD diagnosis (38). Depending on which forms of hypospadias might qualify as DSD, we assume that we have largely underestimated the prevalence of the subgroup of other 46,XY DSD in our study. Interestingly, we observed the highest prevalence in the most recent study period from 2015-2019. Increased awareness of DSD among different specialties may have led to more referrals to pediatric endocrinologists in more recent years, particularly after the Chicago Consensus Statement (2). This group may also include individuals who have not yet received a genetic DSD diagnosis that would allow categorization into the more specific DSD subcategories.

46,XX DSD comprise disorders of gonadal development, androgen excess and other forms. Most studies describe prevalence for individuals with androgen excess, mostly for CAH. Only one nation-wide cohort study from Sweden reported data on other forms of 46,XX DSD (35). They report a prevalence of 3.5-4.7 per 100’000 newborn males for individuals with a 46,XX karyotype and a male phenotype who mainly presented for fertility concerns and were diagnosed with DSD. Among their study cohort of 44 individuals, 33 had disorders of gonadal development and 3 had CAH. Comparison to our data is difficult, because categorization of 46,XX DSD diagnoses differed. In our study, we calculated prevalence for 46,XX DSD based on newborn girls while they used newborn boys, and we did not collect data on phenotype.

Because our cohort was relatively young, we may have missed individuals with 46,XX DSD who only come to medical attention later in life due to fertility concerns, leading to an underestimation of the prevalence in this DSD group. We also assume underreporting of 46,XX disorders of gonadal development and other 46,XX DSD for the earliest interval from 2000-2004 due to the retrospective study design and lack of consensus on diagnostic nomenclature.

Several countries, including Switzerland, published CAH prevalence data from their national neonatal screening programs. Our prevalence estimates are hard to compare to data from the Swiss Newborn Screening (https://www.neoscreening.ch/de), because they do not distinguish between girls and boys and they only capture individuals with classic CAH. Our cohort included individuals with non-classic CAH. Prevalence of all subtypes of CAH in Danish females was 15.1 per 100’000 newborn girls (24), which was comparable to our study with a prevalence of 13.3 per 100’000 newborn girls for 46,XX androgen excess.

Information of population-based cohorts are important to yield reliable estimates on DSD prevalence, and to provide evidence-based data for topics such as sex reassignment, genital surgery, hormonal treatments and other needs of DSD care. In Switzerland, most individuals with any type of DSD born between 2000 and 2019 are cared for by interdisciplinary DSD teams as recommended by international recommendations (1, 4, 8, 10, 39–42). Our study showed that individuals with complex forms of DSD are extremely rare in Switzerland. For these, interdisciplinary care and shared decision-making are crucial, because they require comprehensive diagnostic procedures, and detailed treatment guidelines are lacking. In the last decades, different perspectives from patient representatives, human rights activists, medical professionals and policy makers raised public awareness and led to extensive discussions among stakeholders (43). Controversially discussed is, for example, whether medical treatments offered to children with atypical genitalia who cannot consent should be legally banned and how sex registration should be managed (4, 9). In the multicenter study from Scotland, sex registration was delayed in 9 per 100’000 (1 of 11’097) newborns who had atypical genitalia at birth suggesting a complex form of DSD (23). In our 20-year study period, only 13/543 individuals (2.4 %) had a gender at last follow-up that differed to the registered sex at birth. This indicates that sex registration at birth is mostly fitting. However, we may have underestimated the frequency of gender reassignment in DSD, because data on sex registration were not available in 18 individuals (3.2 %) and gender incongruence might manifest only later in life in our young study cohort.

Our study has some limitations. Data collection covered the whole of Switzerland except for the canton of Ticino. Ticino’s population represents around 4% of the total Swiss population and 3.5% of the newborn population during our study period (25, 44). Ticino has a unique position within Switzerland. It is the only canton where Italian is the official language and is culturally influenced by Italy due to its geographical location in the South of Switzerland and its proximity to Italy. It is unlikely that individuals with DSD born and living in Ticino are treated in other cantons, which allowed us to calculate population-based prevalence of DSD in Switzerland. Our study is also limited by underestimation of prevalence for individuals with DSD. Our time interval of data collection was relatively short and very recent (from 2000-2019), leading to a young study population at time of data collection. Thus, our study did not capture individuals with DSD who have not yet received a diagnosis. Identification of individuals with DSD was retrospective, thus we may have missed patients which were not correctly coded as DSD in the administrative clinical databases or hospital coding systems. As the Chicago Consensus Classification was published and implemented in 2006, and the structure of the I-DSD Registry changed during our study period, classification of DSD might have differed over time. To avoid misclassification, a designated study team coded all DSD cases into categories according to the Chicago Consensus Classification based on extracted data. We could not calculate incidence of DSD, because information on year of diagnosis was missing. We also could not calculate the external genitalia score (EGS) (45) or the external masculinization score (EMS) (46), because the core data set used in this study did not include phenotype information. However, the scope of our study focused on prevalence of DSD according to Chicago Consensus Classification, and we did not require phenotype information for coding.

In summary, we found that individuals with complex forms of DSD were rare and received treatment from interdisciplinary specialized DSD teams. Our data, similar to other studies, underestimated prevalence. National and international registries with complete prospective and standardized follow-up, such as the International Registries For Rare Conditions Affecting Sex Development & Maturation (SDMregistries) (5), are important tools to provide urgently needed evidence-based data. Particularly in the field of rare diseases, international networking is very important. Standard guidelines for analyzing prevalence of DSD are lacking to enable comparison of countries and studies. Such guidelines should include standards on classification of diagnostic groups and a definition of the population at risk for each DSD diagnostic subgroup.

## Acknowledgements

We thank the Swiss DSD Cohort Study Group for their support. Members of this groups who helped in the collection of data are (in alphabetical order): Christine Aebi-Ochsner, Private Practice for Paediatric Endocrinology, Biel Switzerland; Kanetee Busiah, Department of Pediatric Endocrinology and Diabetology, CHUV, University Children’s Hospital, Lausanne, Switzerland; Mirjam Dirlewanger, Pediatric Endocrine and Diabetes Unit, Children’s University Hospital Geneva, Geneva, Switzerland; Sylvia Gschwend, Private Practice for Paediatric Endocrinology, Zug, Switzerland; Melanie Hess, Pediatric Endocrinology and Diabetology, University Children’s Hospital Basel, Basel, Switzerland; Beatrice Kuhlmann, Pediatric Endocrinology, Cantonal Hospital Aarau, Aarau, Switzerland and Private Practice for Paediatric Endocrinology, Basel, Switzerland; Dagmar l’Allemand, Department of Endocrinology, Children’s Hospital of Eastern Switzerland, St. Gallen, Switzerland; Mariarosaria Lang, Paediatric Endocrinology and Diabetology, University Children’s Hospital Zurich, Zurich, Switzerland; Kees Noordam, Centre for Paediatric Endocrinology Zurich (PEZZ), Zurich, Switzerland; Franziska Phan-Hug, Department of Pediatrics, Hospital Morges, Morges and Department of Pediatrics, Hospital Wallis, Sion, Switzerland; Ursina Probst, Department of Pediatrics, Cantonal Hospital Winterthur, Winterthur, Switzerland; Maristella Santi, Department of Pediatrics, Cantonal Hospital Lucerne, Lucerne, Switzerland; Silvia Schmid, Private Practice for Paediatric Endocrinology, Dubendorf, Switzerland; Valérie Schwitzgebel, Pediatric Endocrine and Diabetes Unit, Children’s University Hospital Geneva, Geneva, Switzerland; Michael Steigert, Department of Pediatrics, Cantonal Hospital Graubuenden, Chur, Switzerland; Gabor Szinnai, Pediatric Endocrinology and Diabetology, University Children’s Hospital Basel, Basel, Switzerland; Gerald Theintz, Private Practice for Paediatric Endocrinology, Lausanne, Switzerland.

## Data Availability Statement

Data of this study are registered in the I-DSD Registry and can be accessed upon request. I-DSD governance and rules apply, see https://sdmregistries.org/.

## Supplemental material

**Repository:** https://boris-portal.unibe.ch/handle/20.500.12422/33491

**Supplemental Table 1:** Prevalence, average number of cases per year and sex ratio of children identified with DSD in Switzerland, born between 2000-2019, by diagnostic group and 5-year periods.

**Supplemental Figure 1:** Trends in prevalence of identified DSDs in Switzerland between 2000-2019, by DSD diagnostic group according to the Chicago Consensus Classification.

