## Supplemental Table 1; Supplemental Figure 1 for "Prevalence of differences of sex development in children and adolescents in Switzerland from 2000-2019"

Prevalence, average number of cases per year of individuals identified with DSD in Switzerland, born between 2000-2019, by diagnostic group and 5-year period.

| **2000-2004** | | | |
| --- | --- | --- | --- |
|  | **Average number of cases/year** | **Prevalence per 100.000 newborns per year (95% CI)** | **1:N newborns** |
| *All DSD diagnoses^a^* | 29.2 | 41.3 (34.8; 48.5) | 146 |
| *DSD diagnostic group* |  |  |  |
| Sex chromosome DSD^a^ | 17.0 | 24.0 (19.2; 29.7) | 85 |
| 46,XY DSD^a^ | 7.4 | 10.5 (7.4; 14.4) | 37 |
| 46,XX DSD^a^ | 4.8 | 6.8 (4.3; 10.1) | 24 |
| *Sex chromosome DSD* |  |  |  |
| Klinefelter syndrome and variants^b^ | 6.4 | 17.6 (12.0; 24.8) | 32 |
| Turner syndrome and variants^c^ | 8.4 | 24.4 (17.6; 33.0) | 42 |
| 45,X/46,XY and variants^a^ | 2.0 | 2.8 (1.3; 5.2) | 10 |
| Other^a^ | 0.2 | 0.3 (0.0; 1.6) | 1 |
| *46,XY DSD* |  |  |  |
| Disorders of gonadal development^b^ | 1.8 | 4.9 (2.3; 9.4) | 9 |
| Disorders in androgen synthesis or action^b^ | 2.8 | 7.7 (4.2; 12.9) | 14 |
| Other^b^ | 2.8 | 7.7 (4.2; 12.9) | 14 |
| *46,XX DSD* |  |  |  |
| Disorders of gonadal development^c^ | 0.4 | 1.2 (0.1; 4.2) | 2 |
| Androgen excess^c^ | 4.4 | 12.8 (8.0; 19.4) | 22 |
| Other^c^ | 0 |  |  |
| **2005-2009** | | | |
|  | **Average number of cases/year** | **Prevalence per 100.000 newborns per year (95% CI)** | **1:N newborns** |
| *All DSD diagnoses^a^* | 32.0 | 44.3 (37.7; 51.7) | 160 |
| *DSD diagnostic group* |  |  |  |
| Sex chromosome DSD^a^ | 16.8 | 23.2 (18.5; 28.8) | 84 |
| 46,XY DSD^a^ | 7.8 | 10.8 (7.7; 14.7) | 39 |
| 46,XX DSD^a^ | 7.4 | 10.2 (7.2; 14.1) | 37 |
| *Sex chromosome DSD* |  |  |  |
| Klinefelter syndrome and variants^b^ | 4.0 | 10.7 (6.6; 16.6) | 20 |
| Turner syndrome and variants^c^ | 10.0 | 28.5 (21.2; 37.6) | 50 |
| 45,X/46,XY and variants^a^ | 1.4 | 1.9 (0.8; 4.0) | 7 |
| Other^a^ | 1.4 | 1.9 (0.8; 4.0) | 7 |
| *46,XY DSD* |  |  |  |
| Disorders of gonadal development^b^ | 1.8 | 4.8 (2.2; 9.2) | 9 |
| Disorders in androgen synthesis or action^b^ | 1.2 | 3.2 (1.2; 7.0) | 6 |
| Other^b^ | 4.8 | 12.9 (8.3; 19.2) | 24 |
| *46,XX DSD* |  |  |  |
| Disorders of gonadal development^c^ | 0.4 | 1.1 (0.1; 4.2) | 2 |
| Androgen excess^c^ | 6.8 | 19.4 (13.4; 27.1) | 34 |
| Other^c^ | 0.2 | 0.6 (0.0; 3.2) | 1 |

**Supplemental Table 1 continued**

Prevalence, average number of cases per year of individuals identified with DSD in Switzerland, born between 2000-2019, by diagnostic group and 5-year period.

| **2010-2014** | | | |
| --- | --- | --- | --- |
|  | **Average number of cases/year** | **Prevalence per 100.000 newborns per year (95% CI)** | **1:N newborns** |
| *All DSD diagnoses^a^* | 27.2 | 34.3 (28.8; 40.5) | 136 |
| *DSD diagnostic group* |  |  |  |
| Sex chromosome DSD^a^ | 12.0 | 15.1 (11.5; 19.5) | 60 |
| 46,XY DSD^a^ | 8.8 | 11.1 (8.1; 14.9) | 44 |
| 46,XX DSD^a^ | 6.4 | 8.1 (5.5; 11.4) | 32 |
| *Sex chromosome DSD* |  |  |  |
| Klinefelter syndrome and variants^b^ | 2.4 | 5.9 (3.0; 10.3) | 12 |
| Turner syndrome and variants^c^ | 7.8 | 20.2 (14.4; 27.7) | 39 |
| 45,X/46,XY and variants^a^ | 1.8 | 2.3 (1.0; 4.3) | 9 |
| Other^a^ | 0 |  |  |
| *46,XY DSD* |  |  |  |
| Disorders of gonadal development^b^ | 1.0 | 2.4 (0.8; 5.7) | 5 |
| Disorders in androgen synthesis or action^b^ | 1.6 | 3.9 (1.7; 7.7) | 8 |
| Other^b^ | 6.2 | 15.2 (10.3; 21.5) | 31 |
| *46,XX DSD* |  |  |  |
| Disorders of gonadal development^c^ | 0.2 | 0.5 (0.0; 2.9) | 1 |
| Androgen excess^c^ | 4.8 | 12.5 (8.0; 18.5) | 24 |
| Other^c^ | 1.4 | 3.6 (1.4; 7.5) | 7 |
| **2015-2019** | | | |
|  | **Average number of cases/year** | **Prevalence per 100.000 newborns per year (95% CI)** | **1:N newborns** |
| *All DSD diagnoses^a^* | 23.8 | 28.2 (23.4; 33.7) | 119 |
| *DSD diagnostic group* |  |  |  |
| Sex chromosome DSD^a^ | 7.4 | 8.8 (6.2; 12.1) | 37 |
| 46,XY DSD^a^ | 11.4 | 13.5 (10.2; 17.5) | 57 |
| 46,XX DSD^a^ | 5.0 | 5.9 (3.8; 8.7) | 25 |
| *Sex chromosome DSD* |  |  |  |
| Klinefelter syndrome and variants^b^ | 2.0 | 4.6 (2.2; 8.5) | 10 |
| Turner syndrome and variants^c^ | 3.6 | 8.8 (5.2; 13.8) | 18 |
| 45,X/46,XY and variants^a^ | 1.4 | 1.7 (0.7; 3.4) | 7 |
| Other^a^ | 0.4 | 0.5 (0.0; 1.7) | 2 |
| *46,XY DSD* |  |  |  |
| Disorders of gonadal development^b^ | 1.0 | 2.3 (0.7; 5.4) | 5 |
| Disorders in androgen synthesis or action^b^ | 1.8 | 4.2 (1.9; 7.9) | 9 |
| Other^b^ | 8.6 | 19.8 (14.4; 26.7) | 43 |
| *46,XX DSD* |  |  |  |
| Disorders of gonadal development^c^ | 0.6 | 1.5 (0.3; 4.3) | 3 |
| Androgen excess^c^ | 3.8 | 9.2 (5.6; 14.4) | 19 |
| Other^c^ | 0.6 | 1.5 (0.3; 4.3) | 3 |

Abbreviations: CI, confidence interval; DSD, differences of sex development.

Footnotes:

^a^Prevalence calculated based on newborns of both sexes.

^b^Prevalence calculated based on newborn boys.

^c^Prevalence calculated based on newborn girls.

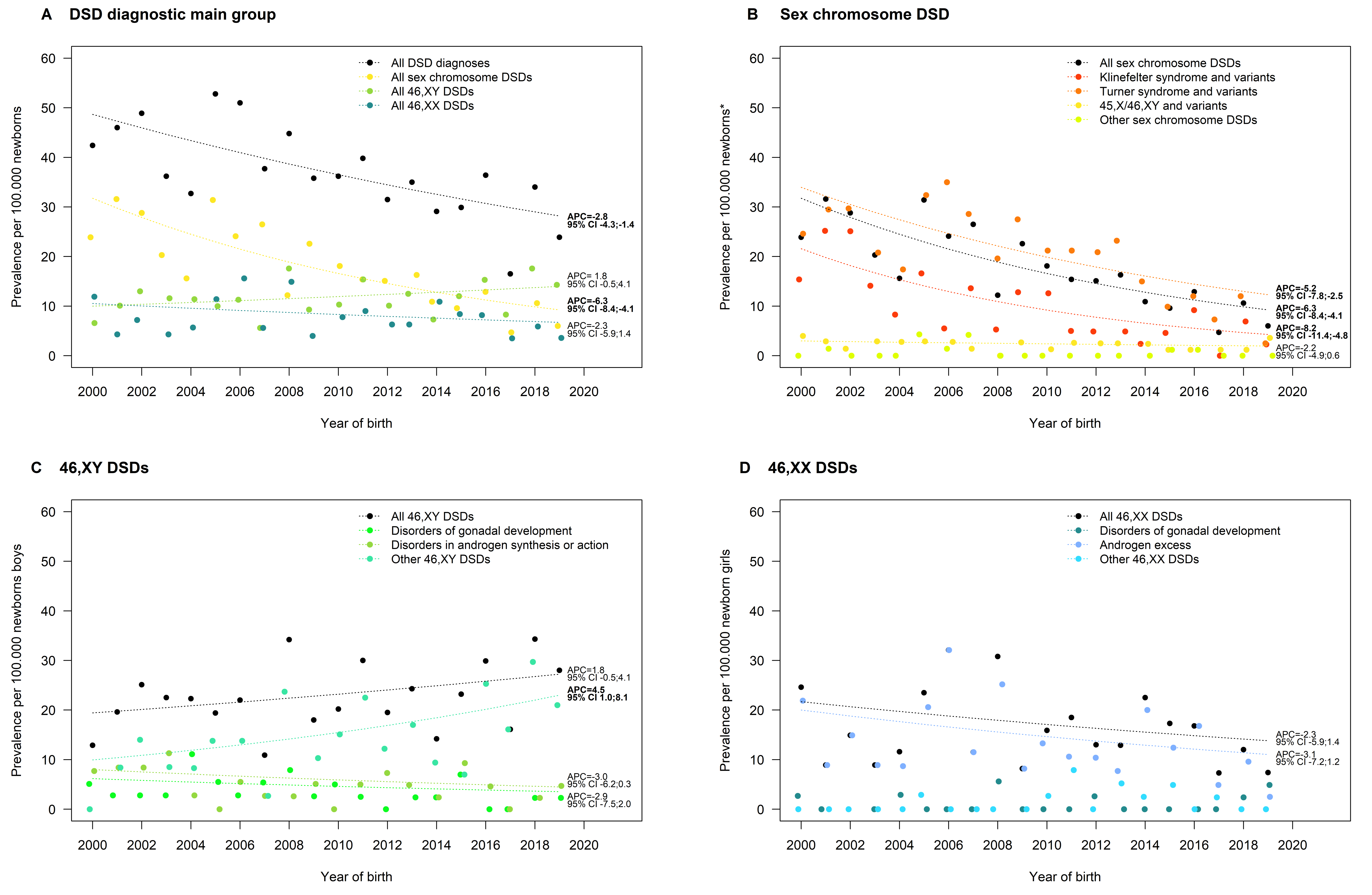

**Supplemental Figure 1:** Trends in prevalence of identified DSDs in Switzerland between 2000-2019, by DSD diagnostic group according to the Chicago Consensus Classification. We used JoinPoint regression to analyze the annual percentage change (APC) in prevalence over time. We did not model APC in diagnostic groups where there were no cases in >3 calendar years. Bold letters indicate p-values <0.5. Abbreviations: APC, Annual Percent Change; DSD, differences of sex development.
